# Exploring the Explainability of a Machine Learning Model for Prostate Cancer: Do Lesions Localize with the Most Important Feature Maps?

**DOI:** 10.1101/2024.10.12.24315347

**Authors:** Destie Provenzano, Shawn Haji-Momenian, Vivek Batheja, Murray Loew

## Abstract

As the use of AI grows in clinical medicine, so does the need for better explainable AI (XAI) methods. Model based XAI methods like GradCAM evaluate the feature maps generated by CNNs to create visual interpretations (like heatmaps) that can be evaluated qualitatively. We propose a simple method utilizing the most important (highest weighted) of these feature maps and evaluating it with the most important clinical feature present on the image to create a quantitative method of evaluating model performance. We created four Residual Neural Networks (ResNets) to identify clinically significant prostate cancer on two datasets (1. segmented prostate image and 2. full cross sectional pelvis image (CSI)) and two model training types (1. transfer learning and 2. from-scratch) and evaluated the models on each. Accuracy and AUC was tested on one final full CSI dataset with the prostate tissue removed as a final test set to confirm results. Accuracy, AUC, and co-localization of prostate lesion centroids with the most important feature map generated for each model was tabulated and compared to co-localization of prostate lesion centroids with a GradCAM heatmap. Prostate lesion centroids co-localized with any model generated through transfer learning ≥97% of the time. Prostate lesion centroids co-localized with the segmented dataset 86 > 96% of the time, but dropped to 10% when segmented model was tested on the full CSI dataset and 21% when model was trained and tested on the full CSI dataset. Lesion centroids co-localized with GradCAM heatmap 98% > 100% on all datasets except for that trained on the segmented dataset and tested on full CSI (73%). Models trained on the full CSI dataset performed well (79% > 89%) when tested on the dataset with prostate tissue removed, but models trained on the segmented dataset did not (50 > 51%). These results suggest that the model trained on the full CSI dataset uses features outside of the prostate to make a conclusion about the model, and that the most important feature map better reflected this result than the GradCAM heatmap. The co-localization of medical region of abnormality with the most important feature map could be a useful quantitative metric for future model explainability.

## 1.0 Background

As the use of AI grows in clinical medicine, so does the need for better explainable AI (XAI) methods.^1 2 3 4 5 6^.^7 8^ Current popular model based XAI methods such as SHapley Additive exPlanations (SHAP), Local Interpretable Model-agnostic Explanations (LIME), and Gradient-Weighted Class Activation Mappings (GradCAM) focus on post-hoc interpretations of the data which can provide unstable or incorrect model explanations.^9 10 11^ A known weakness of these techniques is that they lack quantitative metrics for explainability, and there is a lack of consensus on how to use them to explain model performance.^12 13^ The qualitative nature of these methods can inspire “false hope” in model selections as they force the observer to assess what the model thought was important as part of a post-hoc analysis.^14^ Without quantitative metrics of explainability there remains a gap between a model developer’s urge to explain the model’s inner workings and the clinician’s need to identify clinical relevance of the selections.^15^ This makes quantitative metrics of utmost importance to explain a model’s selections and inspire trust in real-world clinical applications. A study by Jin, et al. proposed the Clinical XAI Guidelines that suggested explainability metrics within medical imaging should meet 5 criteria: [1] understandability, [2] clinical relevance, [3] truthfulness, [4] informative plausibility, and [5] computational efficiency.^16^ In their study, they found that no current popular XAI method met all of these guidelines. In this study, we propose a quantitative model-based XAI method that could satisfy all five criteria.

Model-based XAI methods like GradCAM evaluate the features generated by Convolutional Neural Networks (CNN). The convolutional layer of a CNN identifies important regions of the image by sliding a window (kernel) of predetermined size across the image to produce a weighted “feature map.” Unfortunately, these feature maps are often difficult to interpret as there can be thousands of features for a single input image. GradCAM combines these post-hoc to create a qualitative heatmap interpretation.^17^ GradCAM also suffers from known weaknesses that prevent it from localizing correctly on a target when multiple classes are present or image targets overlap.^18^ We observed in preliminary tests that one of the ways to mitigate these weaknesses was to evaluate feature maps individually, specifically by using the highest-weighted feature map. For this study, we turned the highest-weighted feature map (or “most important” feature map) into a quantitative metric by evaluating whether it contained the anatomic location of an abnormality on Magnetic Resonance Imaging (MRI) data for prostate cancer. Ultimately this study hopes to demonstrate an objective, easy to use, in-model quantitative metric of explainability that does not require extensive post-hoc analyses.

## 2.0 Methods

### 2.1 Data Collection

ProstateX, a publicly available dataset of prostate MRI’s hosted on The Cancer Imaging Archive (TCIA), was used to develop the series of predictive models used for testing in this study.^19^ This dataset consists of 330 prostate MRIs collected at Radboud University Medical Center on a MAGNETOM Trio and Skyra Siemens 3D Magnetic Resonance (MR) scanner, which were initially interpreted by radiologists assessing patients for prostate cancer. Abnormal areas in the gland were labeled as “prostate lesions” and targeted for biopsy. Cancer positive is defined as “clinically significant” (CS) prostate cancer with a Gleason Score ≥ 3+4 (Gleason Grade ≥ 2) while benign “non-clinically significant” (NCS) disease is defined as Gleason score ≤ 3+3 (Gleason grade of ≤ 1). ^20^. This study used T2-Weighted (T2W) images, which were acquired using a turbo spin echo sequence with a 0.5 mm resolution in-plane and slice thickness of 3.6 mm. A random selection of 63 lesions from the NCS cohort was selected to create a balanced dataset with a 50/50 split of 63 CS and 63 NCS lesions. Image slices with more than one lesion on a given image slice were excluded.

### 2.3 Dataset Preparation

This study used the small field of view axial T2 sequence of the lower pelvis and prostate, shown in figure 1. The cancer diagnosis and centroid of each prostate lesion was determined by an abdominal radiologist at Radboud University Medical Center institution. An experienced radiologist from our institution (SHM) segmented prostate image slices to generate three datasets: (1) segmented prostate image, (2) full cross-sectional pelvis image (CSI), and (3) a pelvis image with no prostate tissue (“Prostate Removed”).

**Figure 1:**
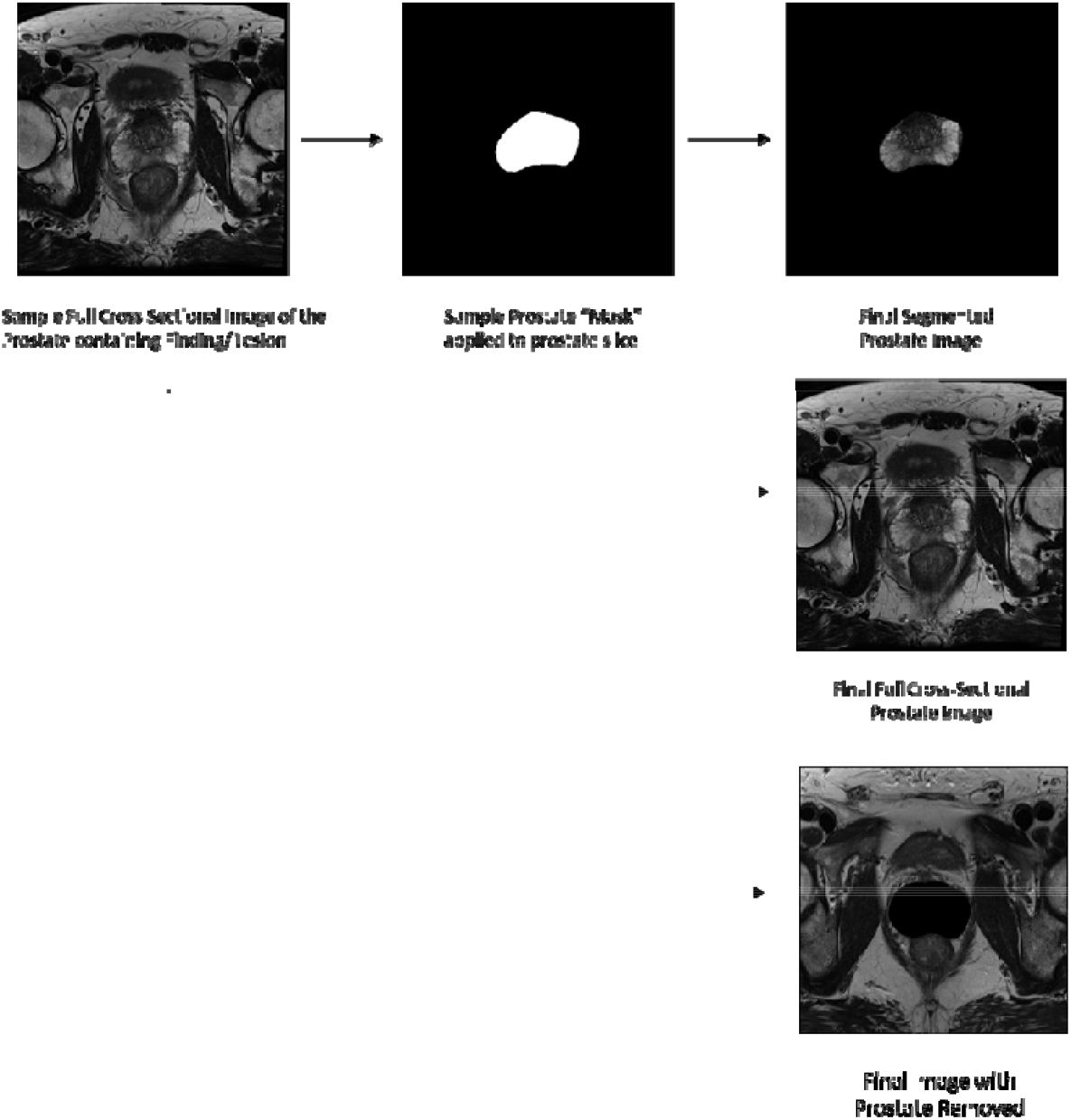
Generation of prostate masks to create segmented dataset by Board Certified radiologist.

### 2.4 Model Training

The Python TensorFlow package was used to generate and train two types of Residual Neural Network (ResNet) models on two of the training datasets (full CSI and segmented images) to create four types of models:

[1] “From-scratch” model using the ResNet framework in which the weights for every layer were re-estimated on the training dataset.
[2] “Transfer learning” model where only the the final fully connected layer of a model trained elsewhere (ImageNet) was retrained on our data.^21^

Models were then tested on [1] the segmented prostate images [2] full CSI pelvis images and [3] an additional test on the full CSI images with the prostate removed.

To ensure statistical significance, we used 5-fold cross validation by dividing the dataset into five different 80/20 training/ testing “folds” in which each fold was used to separately repeat the entire training, testing, and localization process. This testing set utilized the same image slices across the segmented prostate image data and full CSI pelvis image data. Average accuracy and Area Under the Receiver Operating Characteristic ROC Curve (AUC) were computed across the five folds.^22^ A schematic of this study design is represented in Figure 2.

**Figure 2:**
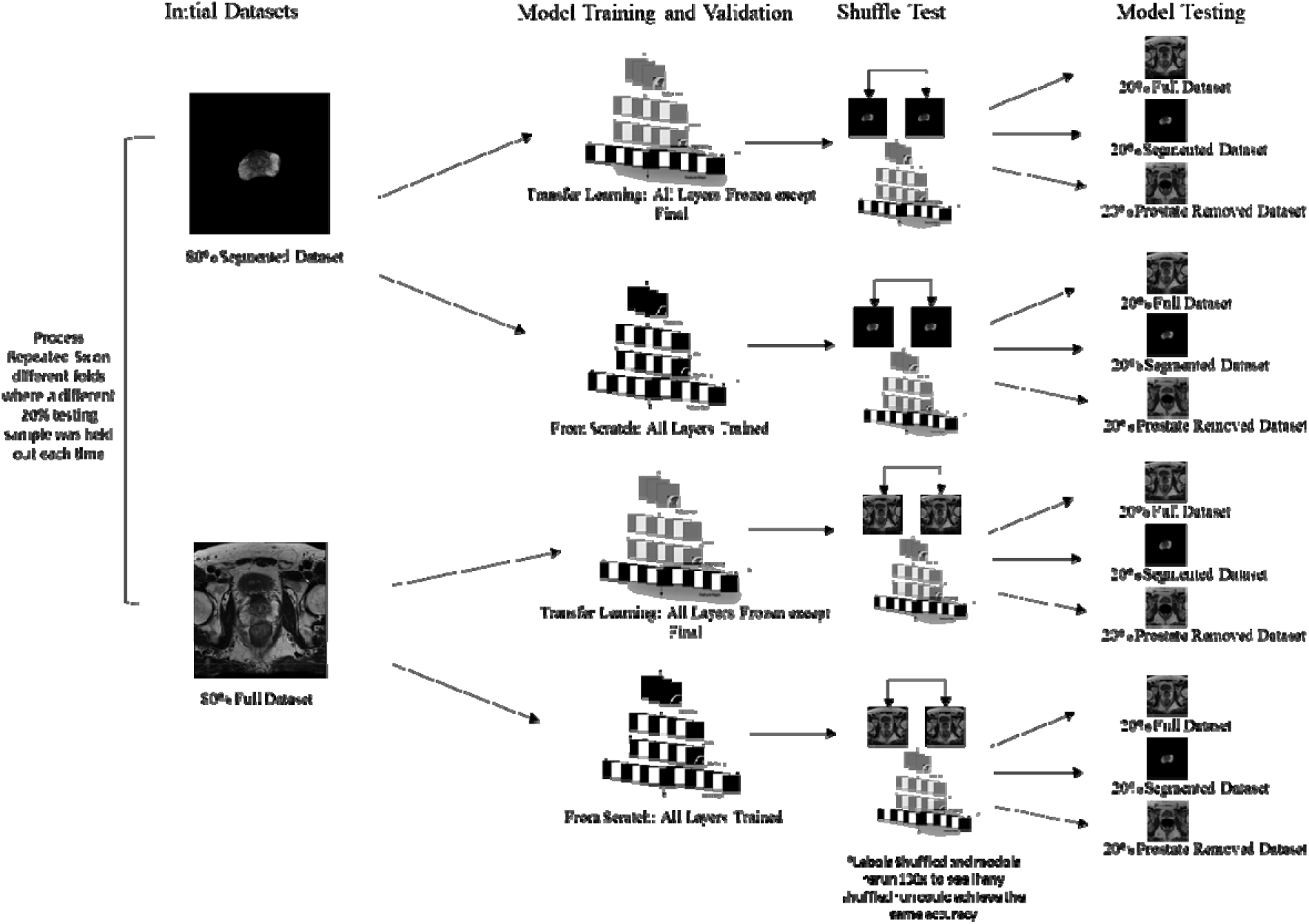
Schematic of full model training and testing combinations including the input dataset (Segmented), two model types (from-scratch, transfer), three final 20% holdout samples (Full CSI pelvis image, segmented prostate image, prostate removed image).

### 2.5 Feature Map Generation and Lesion Localization

Feature Maps were generated on the 20% testing sets by using the final feature-generation layer one step before the sigmoid classification function. In Python packages like TensorFlow or pytorch this can be done for any layer by outputting model predictions after selecting the relevant layer. Feature maps with the most predictive (highest) weights were considered the “most important” for the algorithm. The non-zero regions of the most-important feature maps were tested for localization with the centroids of each prostate lesion.

### 2.6 Statistical Significance and Analysis

Initial models were validated for statistical significance using a “Randomization” (or “Shuffle”) test.^23^ This was done by generating one hundred datasets with shuffled labels to determine whether any randomly shuffled dataset used in the model training process could achieve the same accuracy as the initial model and dataset. It can be assumed that if a randomly shuffled model could achieve the same or greater accuracy as the initial comparison accuracy fewer than five times on one hundred runs, this would correspond to a p < 0.05.

### 2.7 Comparison to Existing Methods

We compared our localization results with Gradient Weighted Class Activation Mappings (GradCAM). GradCAM heatmaps were created for each image using Python TensorFlow.^24^ Localization of the prostate lesion was compared with the maximum value on the GradCAM heatmap, and (2) localization of the prostate lesion with **any** value on the GradCAM heatmap greater than zero. For this second test, any pixel with a GradCAM calculated weight was considered. This method was selected for ease of use and because the qualitative nature of GradCAM makes it difficult to easily analyze without extensive post-hoc analyses. A t-test was used to compare the group averages, the GradCAM results, and the highest-weighted feature map results.

## 3.0 Results

### 3.1 Lesion Localization Compared to Model Performance

Table 1 presents the accuracy and AUC for the model performances for each training and testing run at predicting clinically significant (CS) vs not clinically significant (NCS) cancer. These are the averages from the 5-fold cross validated testing sets. The percentage of lesion centroids that localized with (1) the most important feature map and (2) any non-zero region of the GradCAM heatmap for the corresponding prostate image and full CSI pelvis image are also reported below. **No Lesions correctly localized** with the maximum pixel value (“brightest”) region of the GradCAM heat map. Example images are displayed in Figure 1.

**Table 2:**
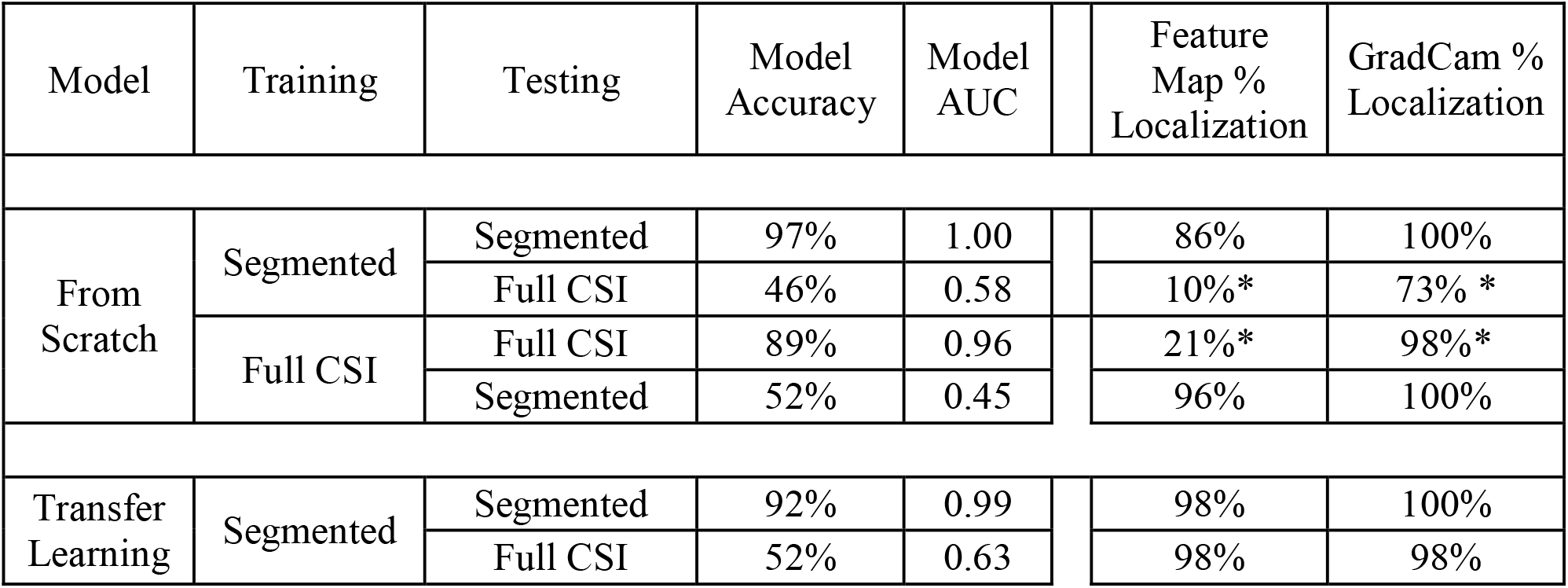
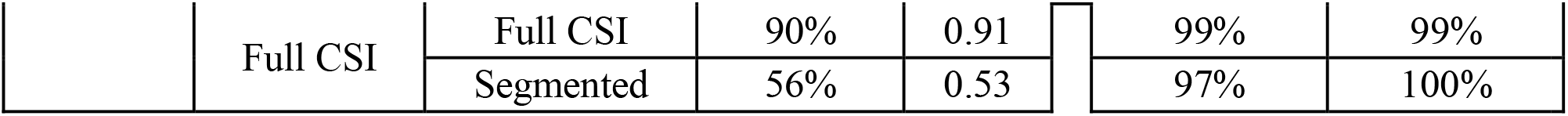
Average Accuracy and AUC across each of the 5-fold cross validated models for model prediction of clinically significant (CS) vs not clinically significant (NCS) prostate cancer and percentage of prostate lesions localized with the most important feature map and any non-zero region of the GradCAM heatmap 1. p < 0.001 for all model results according to Shuffle test. 2. Additional t-test was used to determine whether means of 5-fold cross validated runs significantly differed between corresponding testing sets (Segmented vs Full CSI model) for a given model type and training data combination (for example the corresponding segmented vs full CSI testing data was compared for the model trained on Segmented data using the from scratch technique) All showed a statistically significantly different average at the p 0.05 level. 3. Group means were compared by t-test to means from the most important feature map. *Indicates localization result for GradCAM heatmap statistically significantly differed by t-test from that of the most important feature map for the 5-fold cross validated localization results, p < 0.05

**Figure 1:**
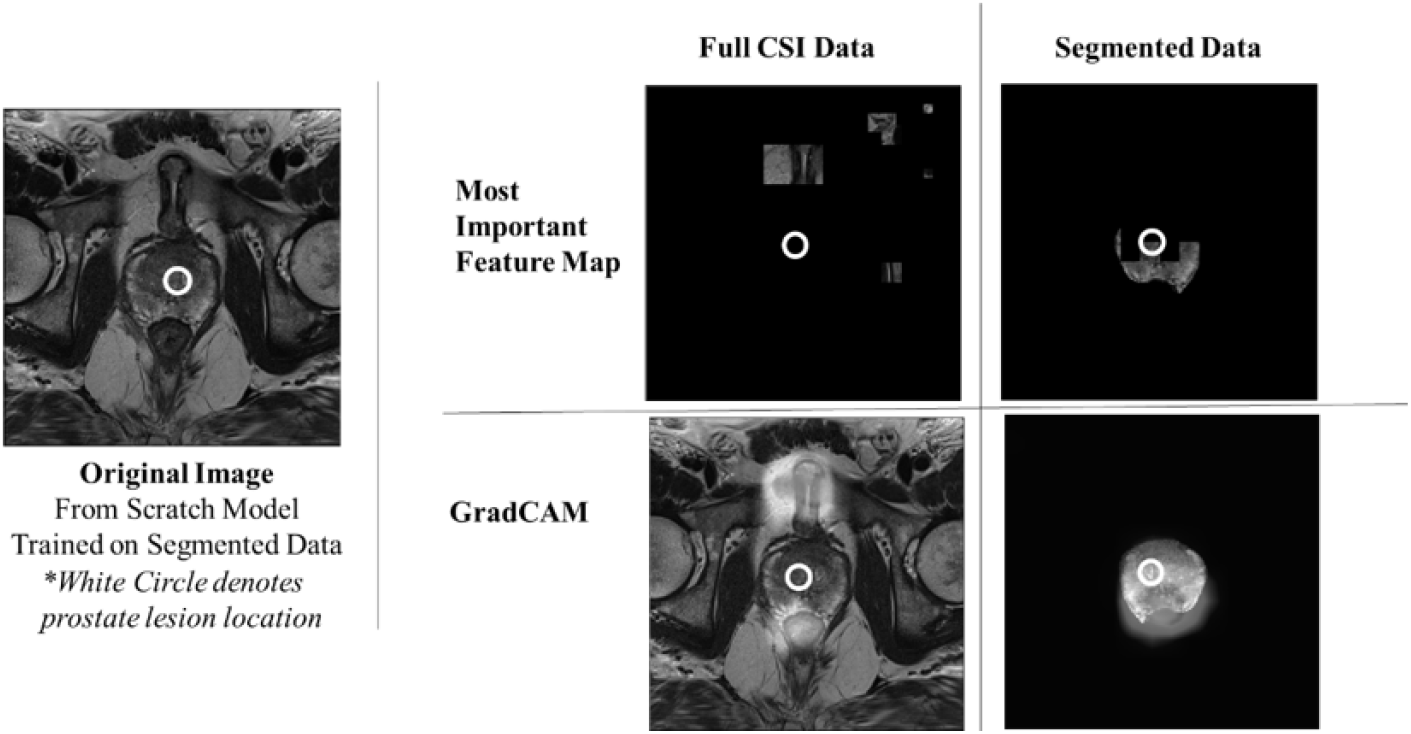
Example most important feature maps compared to GradCAM heatmaps for full CSI and segmented image data. White circle denotes prostate lesion localization. On the GradCAM heatmap, the lighter overlay corresponds to the regions of more importance. More sample feature maps are included in the supplement (Figure S1)

The most important feature maps from the “from scratch” model exhibited different localization when trained and tested on different imaging types (trained segmented -> segmented 86%, full CSI 10%) (trained full CSI 21% -> segmented 96%). The most important feature maps from the models trained using transfer learning localized well (<u>≥</u>97%) with the lesion regardless of testing dataset.

### 3.2 Accuracy on Dataset with Prostate Removed

The initial results suggested that the model trained from scratch on the full CSI data was using patterns in the image outside the prostate, which prompted an additional test on a dataset with prostate tissue removed. Accuracy and AUC for this final testing dataset are reported in Table 3.

**Table 3:** Model Accuracy and AUC on dataset with prostate removed for each model training type (from scratch or transfer learning) and each training dataset (segmented or full CSI). No new models were trained for this final experiment on the new testing set.

| Model | Training | Testing | Model Accuracy | Model AUC |
| --- | --- | --- | --- | --- |
| From Scratch | Segmented | Prostate Removed | 50% | 0.47 |
|  | Full CSI | Prostate Removed | 79% | 0.93 |
| Transfer Learning | Segmented | Prostate Removed | 51% | 0.52 |
|  | Full CSI | Prostate Removed | 88% | 0.89 |

The model trained from scratch on the full CSI dataset maintained a high accuracy/ AUC (79%/ 0.93) even with the prostate removed. Additionally, the model trained using transfer learning on the full CSI dataset was able to maintain a high accuracy and AUC (88%/ 0.89) on the dataset with the prostate removed.

## 4.0 Discussion

All models were able to statistically significantly predict CS vs NCS classification status when trained and tested on similar types of images, regardless of field of view. Models remained highly accurate when training and testing on the full CSI images, which included extraneous data outside the prostate. Performance uniformly dropped when models were trained and tested on different image sets (ie segmented -> full CSI) The additional image data or loss of image data significantly impacted the model resulting in decreased accuracy. These findings were concordant with other studies which have found that model generalizability, or replicability, is one of the known problems facing clinical application of current AI methods.^25 26^

Both models showed high localization with both GradCAM and the most important feature map when models were trained and tested on segmented images. While this initially seems to suggest both models are localizing well, additional testing contradicts this and illustrates very interesting additional caveats. GradCAM showed high localization (73%) and the most important feature map reported poor localization (10%) when the segmented from scratch model was tested on the full CSI data. Given the poor accuracy of the segmented model on the full CSI data and prostate-removed data, the most important feature map localization appears to reflect that additional image data from outside the prostate impacts the model. The models trained using transfer learning reported high localization for both GradCAM and the most important feature map. When model weights previously assigned to features in the prostate are incorrectly assigned to features outside the prostate, the model naturally does not come to the correct conclusion.

Both types of models showed high accuracy when training and testing on the full CSI images. The full CSI model trained from scratch had high localization with GradCAM and poor localization with the most important feature map. Once again, the most important feature map presented a better assessment of localization as highlighted by the results when the prostate was removed. In this scenario, model accuracy remains high once the prostate is removed, illustrating that the model is relying on image data outside of the prostate for classification. Both types of full CSI trained models report poor accuracy but high localization on the segmented images. This implies that when model weights applied to features outside the prostate are then applied to features inside the prostate, the model is not able to come to the correct conclusion. This was confirmed by the high accuracy found when testing on the data with the prostate removed which showed that the features outside the prostate produce accurate classification decisions.

Models built using transfer learning localized with the prostate lesion regardless of use of full CSI pelvis image or segmented prostate image datasets. This explainability analysis confirms the advantage for transfer learning on small training datasets is independent of segmentation. This result was expected, as models trained using transfer learning use “features” from an unrelated previously trained model to better create a new model. The initial potential features generated by a CNN for any image will be the same across any model type with the same initial model parameters, but are weighted differently into potential feature maps by the model training process. Models trained using a transfer learning process have a much larger number of potential features fed to the final fully connected layer as they are generated from a much larger initial dataset (ImageNet) than the from scratch models. But this means they also contain more extraneous data including that outside the prostate (as showcased by the high accuracy maintained on the prostate removed dataset). These results suggest that additional considerations, such as the false positive rate or whether exterior defining characteristics are also included in the feature map, must be considered in addition to the feature map localization. Further studies are recommended to fully explore these possibilities.

There are many additional avenues one could take the results of this study. For example, one could expand the search area from looking to see whether a prostate lesion is contained within a feature map, to looking to see whether <u>any</u> area inside the prostate at all is considered. It’s possible that additional layers within the model may be more illustrative than the final one. Alternatively, a threshold and combination strategy could be used. Three of the 5 criteria defined by Jin et. Al were satisfied in both their study as well as our method: [1] understandability, [2] clinical relevance, and [3] computational efficiency, however no current explainability metric was able to satisfy the last two following conditions: [1] truthfulness, where the explanation matches the model prediction, and [2] informative plausibility where human evaluation of the explanation can reveal model prediction quality. This new feature map metric proposed here provides a potential way to satisfy both, as [1] the most-important feature map directly reflects the model prediction directly and [2] a human could pick a localization that corresponds with the area of known pathologic disease based on absolute truth (biopsy results). Future work with more clinical data would help to evaluate these criteria.

This methodology is not without limitations. The use of only one feature map means there will be inevitably be a loss of data. It is possible that a region that may be important in the top five feature maps may be just as important as the top feature map, which in the current methodology would be overlooked without additional analysis. This analysis benefited from a specific coding framework to generate the most important feature map as it used Python TensorFlow. Models trained using a different framework may require more analysis and code to determine the highest weighted feature map. The dataset used to train and test these results was also ultimately very small. This study would benefit from more data to test the clinical utility of the most important feature map.

The implementation and utility of predictive models in a real-world setting will depend on its accuracy as well as explainability. Some form of testing will be necessary to ensure a model is using “appropriate logic” to arrive at its predictions. Without this explainability testing, there is a risk the model could identify a random transient predictive pattern that could pose a risk of poor generalizability and failure on additional data. The metric proposed in this study provides an easy way to ensure the model is looking at the correct region on an image in a quantitative fashion.

## 5.0 Conclusion

The current state of machine learning research produces models that can accurately predict something has cancer, but these accurate models cannot subsequently find the cancer or produce clear criteria for why it was selected. The location of the cancer and criteria for selection is of utmost importance for a Radiologist. This study evaluated a series of highly accurate models that do not generalize well to a subsequent modified dataset (Segmented -> Full CSI or Full CSI -> Segmented). For the models considered in this study, the inability of a model to localize either on the (1) initial training dataset or (2) final testing dataset combined with a high false positive rate present on the most important feature map offers a partial explanation as to why the models do not generalize well, as they are unable to find the cancer. Additionally, although it is interesting that one can build a highly accurate prostate cancer model that does not use the prostate cancer to make its prediction, such a model would be ineffective in a real-world setting. One should consider metrics such as the localization rate of the prostate lesion with the most important feature map while building a predictive model to ensure the model is useful for a clinician. Additional considerations need to be considered for a transfer learning model beyond the scope of this study to find ways to ensure it is useful at predicting cancer and finding the subsequent cancer. This methodology presents a potential way to validate whether a predictive model trained on clinically significant prostate cancer is “looking” at the anatomical regions of relevance: the prostate lesion. This percentage of targets localized with the most important feature map could be a useful quantitative metric for future studies as well.

## Data Availability

The Cancer Imaging Archive (TCIA) ProstateX collection: https://www.cancerimagingarchive.net/collection/prostatex/

## Abbreviations

ML: Machine Learning
ResNet: Residual Neural Network
DL: Deep Learning
pCA: Prostate Cancer
CS: Clinically Significant Prostate Cancer
NCS: Not Clinically Significant Prostate Cancer
CSI: Cross Sectional Image

## 6.0 Supplement

**Figure S1:**
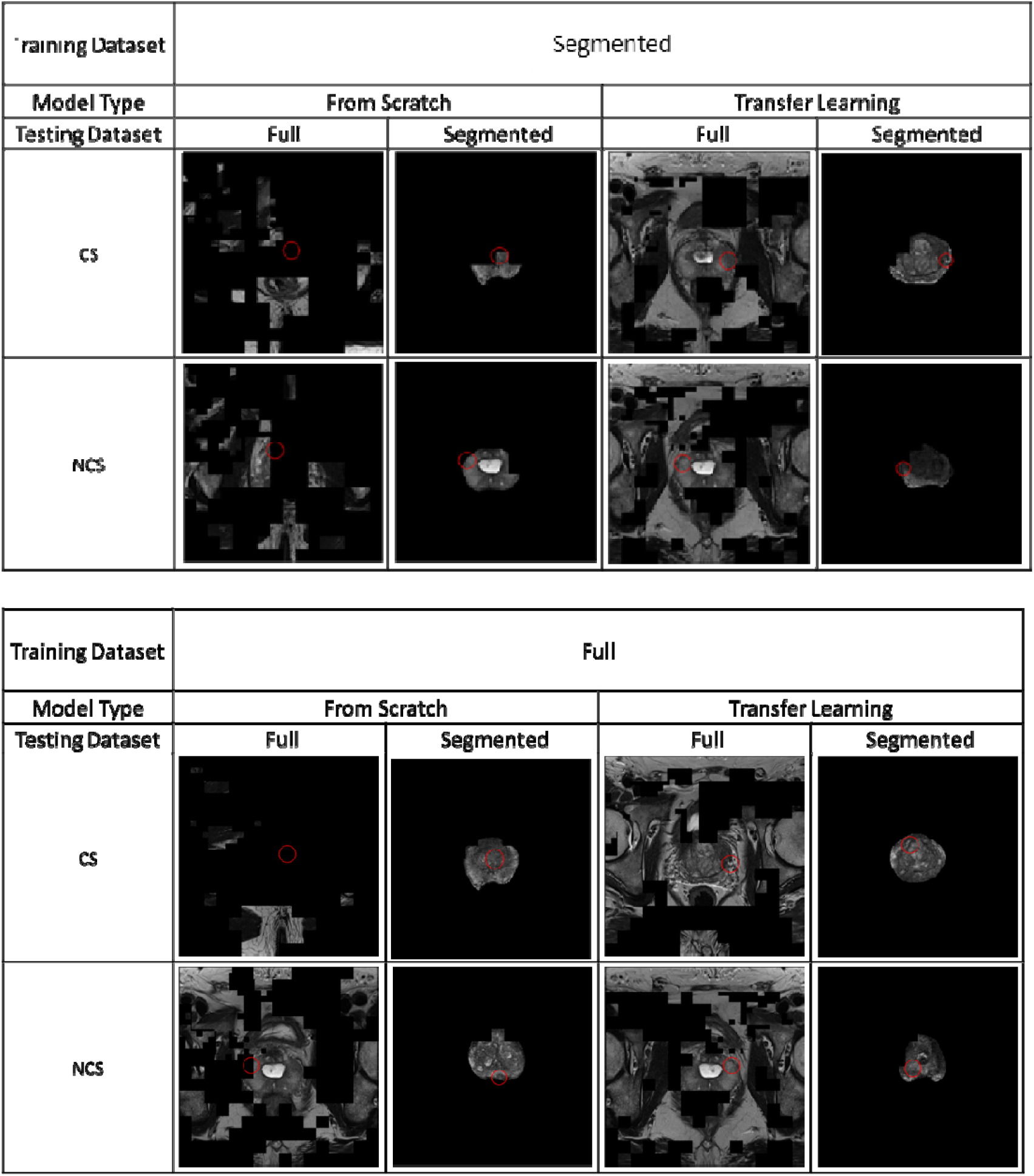
Sample Feature Maps.

